# Use of therapeutic play in nursing care for school-age children: a scoping review

**DOI:** 10.1101/2022.03.18.22272614

**Authors:** Luanna Gomes da Silva, Célida Juliana de Oliveira, Joseph Dimas de Oliveira, Paulo Felipe Ribeiro Bandeira, José Hiago Feitosa de Matos, Emiliana Bezerra Gomes, Izabel Cristina Santiago Lemos de Beltrão, Álissan Karine Lima Martins

**Affiliations:** Programa de Pós-Graduação em Enfermagem, Universidade Regional do Cariri, Crato, Ceará, Brazil; Programa de Pós-Graduação em Enfermagem, Departamento de Enfermagem, Universidade Regional do Cariri, Crato, Ceará, Brazil; Departamento de Enfermagem, Universidade Regional do Cariri, Crato, Ceará, Brazil; Universidade Regional do Cariri, Crato, Ceará, Brazil: Universidade Federal do Vale do São Francisco - UNIVASF

**Author notes:** Correspondence author: Paulo Felipe Ribeiro Bandeira. Universidade Regional do Cariri. Rua Cel. Antônio Luís 1161, Pimenta, 63.100-000, Crato, Ceará, Brazil.

**Keywords:** Child, Play and Playthings, Pediatric Nursing

## Abstract

**Background:** Therapeutic play is an important strategy in nursing care to relieve emotional discomfort of school-age children. The aim of this scoping review was to map the use of therapeutic play in nursing care for school-age children.

**Methods:** *S*earches in the databases: MEDLINE, CINAHL, Scopus, BDENF, LILACS and CAPES theses and dissertations portal. A total of 1,486 records were identified and 32 included in the review. Eligibility criteria: The literature on children aged 6 to 12 participating in therapeutic play sessions in nursing care in any place, in the form of original articles, theses and dissertations, in English, Spanish or Portuguese. There was no time limit.

**Results:** The focus theme has been described since 1987 in the national and international scientific literature. There is consensus on the benefits of therapeutic play in nursing care for school-age children in various situations, especially in the hospital context, with proven efficacy in research that encourages the application of this strategy in nursing practice, but there are barriers for its implementation.

**Conclusion:** The review identified the settings, situations, benefits, barriers and gaps regarding the use of therapeutic play in nursing care for school-age children.

**Implications:** The positive results reported in the studies can be used to enrich discussions about the use of therapeutic play in nursing practice for school-age children and encourage awareness of the importance of this strategy. The gaps identified contribute to future research.

## Introduction

Nursing’s role in child health care allows the integration of specific knowledge to ensure the quality of care and skills that provide good conditions of interaction with the child (Brondani & Pedro, 2019). One of the resources that favors this interaction is play, an essential activity for the development and well-being of the child that does not cease when falls ill. When the opportunity to play is not offered, it may present behavioral changes, such as irritable mood, aggressive attitudes, difficulty in social interaction and developmental delay (Oliveira et al., 2015).

One aspect of the modalities of play is therapeutic play (TP), a care strategy commonly used to relieve children’s anxiety and promote psychosocial well-being (Burns-Nader & Hernandez-Reif, 2014; R. D.M. Silva et al., 2017). Among the modalities of TP was included the instructional (ITP), which prepares for health procedures to which the child will be submitted; the dramatic (DTP), which allows the emotional discharge of the child including the manifestation of feelings, desires and experiences; and the enabler (ETP) of physiological functions, potentiating the rehabilitation of physiological functions in cases where the child shows regressive behavior for the age (Lemos et al., 2016a; Pennafort et al., 2018).

The TP is a structured toy, applied by a professional trained in the technique and with toys recommended in the literature, including cloth or plastic dolls, puppets, animals, carts, graphic material, real or symbolic materials representing the hospital and domestic settings (Maia et al, 2020).

The practice of TP has been described in the literature as an important strategy in nursing care for children in different health settings because it is a resource that favors the provision of holistic and humanized care due to its numerous benefits (Costa et al., 2016; Gesteira et al., 2011; Santos et al., 2017). For the nursing team, it serves as a strategy to establish communication with the child, prepare them for procedures, know their feelings and concerns. For the child, it helps to express their perceptions and needs, alleviate fears, tension and anxiety generated by experiences that seem threatening, optimizing integral well-being and reducing harm in child development (Caleffi et al., 2016; Pennafort et al., 2018).

Studies indicate that children at school-age are still vulnerable to emotional discomfort arising from the disease process, which reduce their sense of control and power, favoring the emotional discomfort caused by the experience of illness, such as fear, anguish and sadness (Nóbrega et al., 2010; S. G. T. Silva et al., 2017). Therefore, the school-age children need to receive assistance that allows promoting their emotional well-being. Nursing professionals have a relevant role to listen to, believe and meet the emotional needs expressed by this child through therapeutic play, which is a child’s right, and should be incorporated into nursing care (Costa et al., 2016; Nóbrega et al., 2010; World Health Organization Regional Office for Europe, 2017). Thus, it is not enough to have a defined place for recreational games; it is necessary to use TP aiming to achieve all the therapeutic benefits it provides (Costa et al., 2016; Costa et al., 2019).

The World Health Organization (WHO), with the objective of guaranteeing children effective health care based on best practices, addresses the need to offer children emotional support, reiterating the right of children to play therapeutically in health units (World Health Organization, 2018). It is worth mentioning the relevance of recreational activities for children, is assured by the United Nations Children’ Fund (1990).

However, studies show that even in view of the importance and recommendation to incorporate TP in pediatric nursing care, its practice is still modest or not observed in the routine of the teams, encountering difficulties regarding its use (Costa et al., 2016; Costa et al., 2019; Francischinelli et al., 2012; Santos et al., 2020). Moreover, there are several studies developed on the effects of TP in all phases throughout childhood, however, a broad survey of studies addressing the practice of TP in nursing care for school-age children still represents a gap in the literature.

In view of these aspects, the need to perform a scoping review is justified to explore the knowledge available in the literature about the use of TP in nursing care for school-age children with emphasis on its application in the care process, the benefits for care and the barriers for its implementation. Thus, the aim of this scoping review was to map the use of therapeutic play in nursing care for school-age children.

## Methods

### Design

This scoping review was undertaken following the JBI methodology (Peters et al., 2020) and PRISMA Extension for Scope Reviews (PRISMA-ScR): Checklist (Tricco et al., 2018). This type of review is a form of knowledge synthesis, addresses a broad research issue and allows to provide an overview of a topic, in addition to mapping the evidence, clarifying research areas and identifying gaps for future research (Peters et al., 2020).

The strategy guided by the mnemonic Population, Concept and Context (PCC) based on the construction of the research question: “What is the state of the art on the use of therapeutic play in nursing care for school-age children?”, the following elements being: P-school-age children; C-therapeutic play; C-nursing care.

### Eligibility criteria

The inclusion criteria were formulated to allow obtaining the largest number of publications available. In relation to the population, studies were included in which children aged 6 to 12 participated in TP sessions, according to school-age defined by reference literature in pediatric nursing (Hockenberry & Wilson, 2011). In the concept, studies were included with TP applied as a structured toy, including cloth or plastic dolls, puppets, animals, carts, graphic material, real or symbolic materials representing the hospital and domestic settings (Maia et al., 2020). Regarding the context, studies were included with TP applied by nursing in any nursing care area.

This review included empirical studies, theses and dissertations, published in English, Spanish or Portuguese. There was no time limit. This review excluded duplicate studies, with full text unavailable in open access and for not meeting the objective of this study.

### Information sources

The search strategy for scientific production was carried out between June and July 2020, in three stages, by two independent reviewers. In the first, a broad search was conducted, with the descriptors child [MeSH] and nursing [MeSH] and therapeutic play [keyword] limited to the CINAHL (Ebsco) and MEDLINE (PubMed) databases, with the purpose of analyzing the title words, abstract and descriptors of the studies, and the keywords and descriptors that aligned with the objective of this study were removed to assemble the search keys of the next step.

In the second stage, using the search keys elaborated, new searches were performed in the databases: MEDLINE, CINAHL, Scopus, LILACS and BDENF. It is notelike that the search keys were composed of descriptors MeSH and DeCS (LILACS and BDENF) and keywords, interconnected by boolean “AND” or “OR” operators.

In the third stage, all the articles included in the review, after reading in full, had their list of references explored in search of additional studies that had not been mapped in the previous stages.

As for the grey literature (Peters et al., 2020), it was researched in May 2021 in the database of theses and dissertations of CAPES (Coordination for the Improvement of Higher Education Personnel), using “child” and “therapeutic play” and “nursing”.

Table 1 describes the search strategy adopted in each database and the identified studies.

**Table 1.**
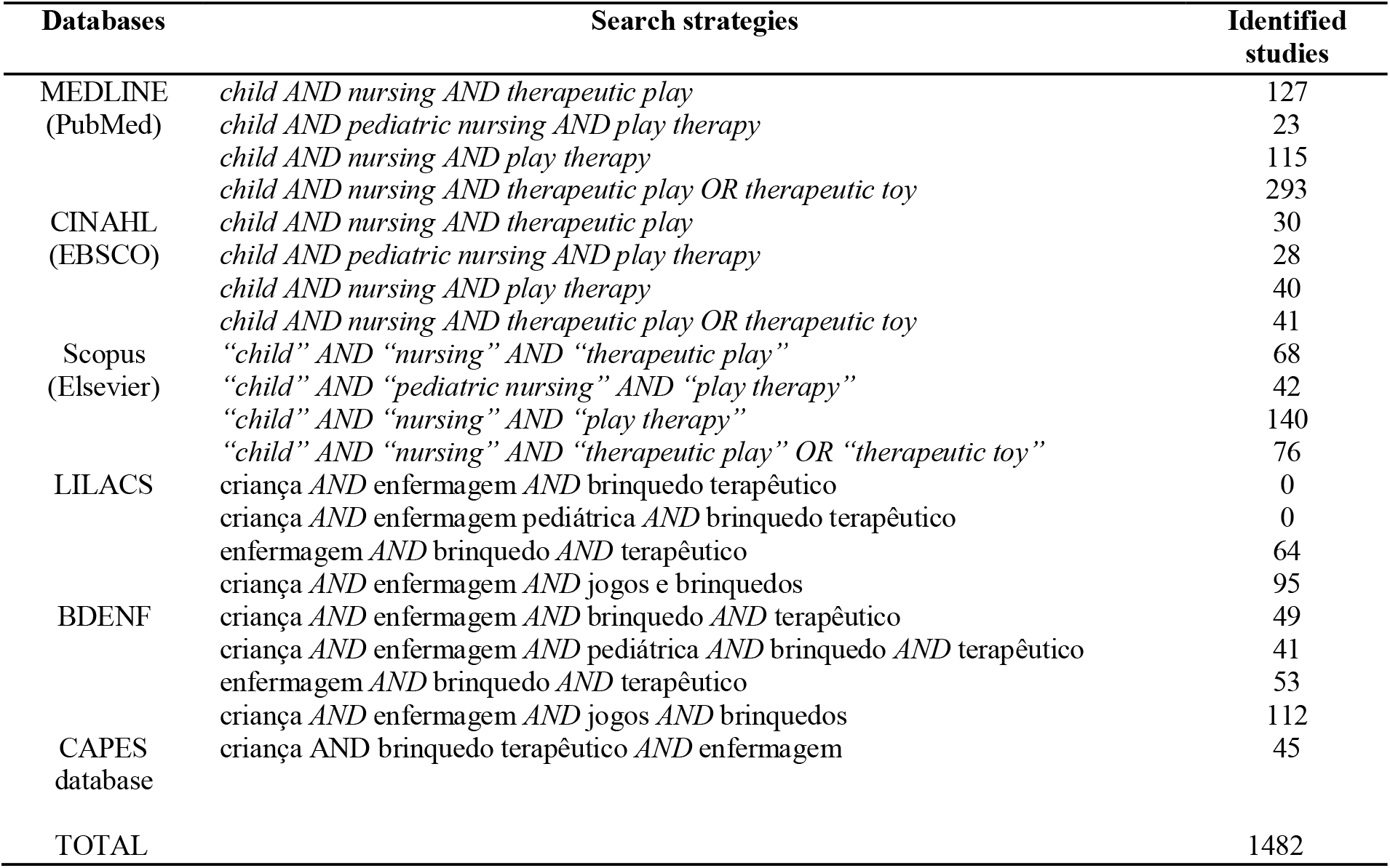
Databases, search strategies and identified studies.

### Selection sources of evidence

Two reviewers participated in the selection of studies, based on the pre-specified inclusion criteria. The divergence regarding the inclusion of some study was treated with a third reviewer.

### Data extraction and analysis

The data extraction of the studies used the standardized JBI data extraction tool (Peters et al., 2020), which made it possible to organize the data based on the characterization of the studies, details about the population, concept and context, and main findings related to the questions of the review, and descriptive analysis was performed.

## Results

From the 1,482 studies, duplicates (n=823) were excluded, which did not meet the inclusion criteria by reading the title (n=431) and abstract (n=99). Subsequently, 133 studies were read in full, being excluded because they did not meet the objective of the review (n=88) and full text unavailable in free access (n=13), and 28 studies were selected that, after analyzing their references, allowed the identification of four more studies. The final sample consisted of 32 studies (Fig.1).

**Fig. 1.**
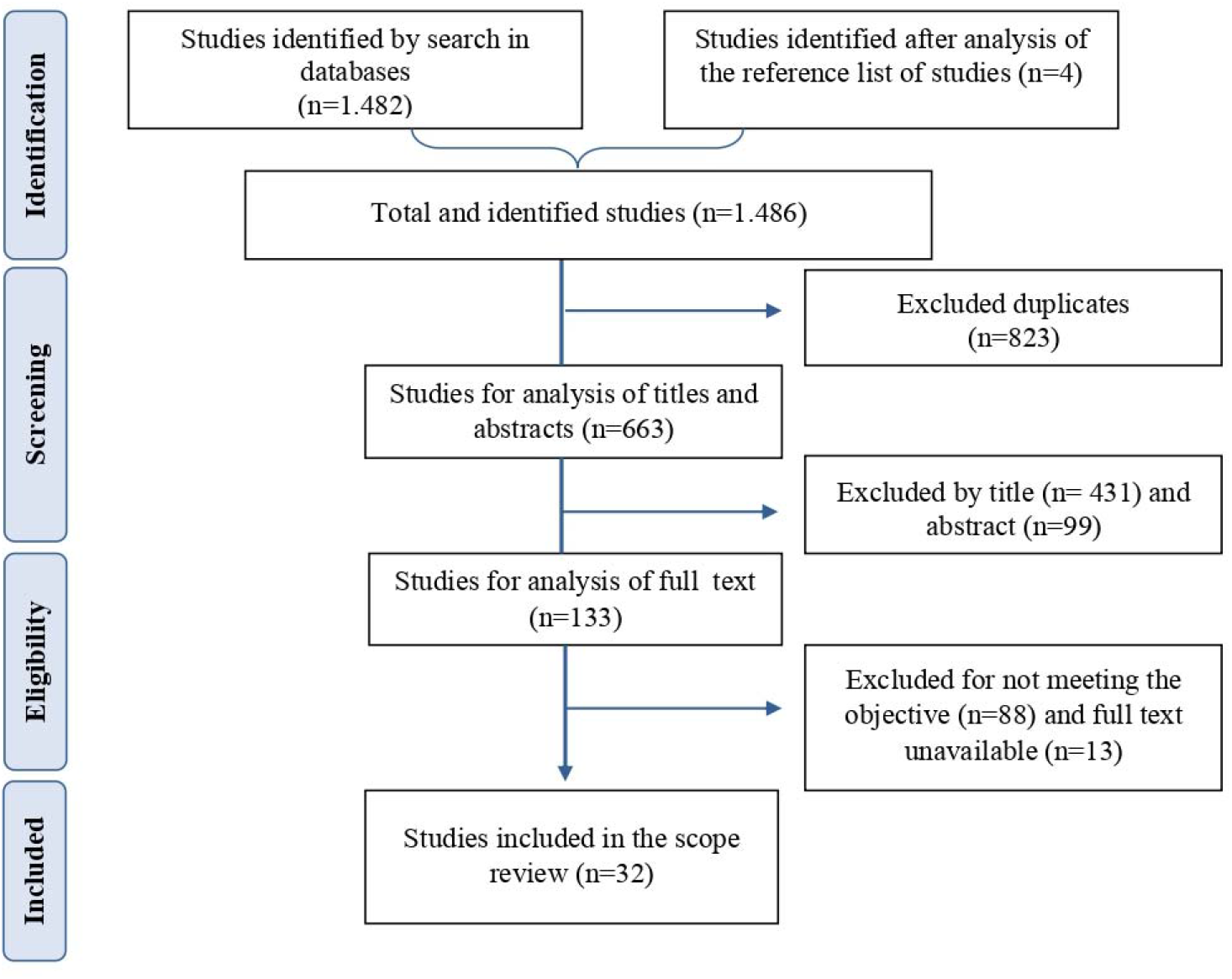
Flowchart of the study selection process according to PRISMA-ScR and JBI recommendations. Source: adapted from Moher et al. (2009).

Table 2 presents the information that characterizes the 32 studies included in the sample analyzed in this scoping review.

**Table 2.**
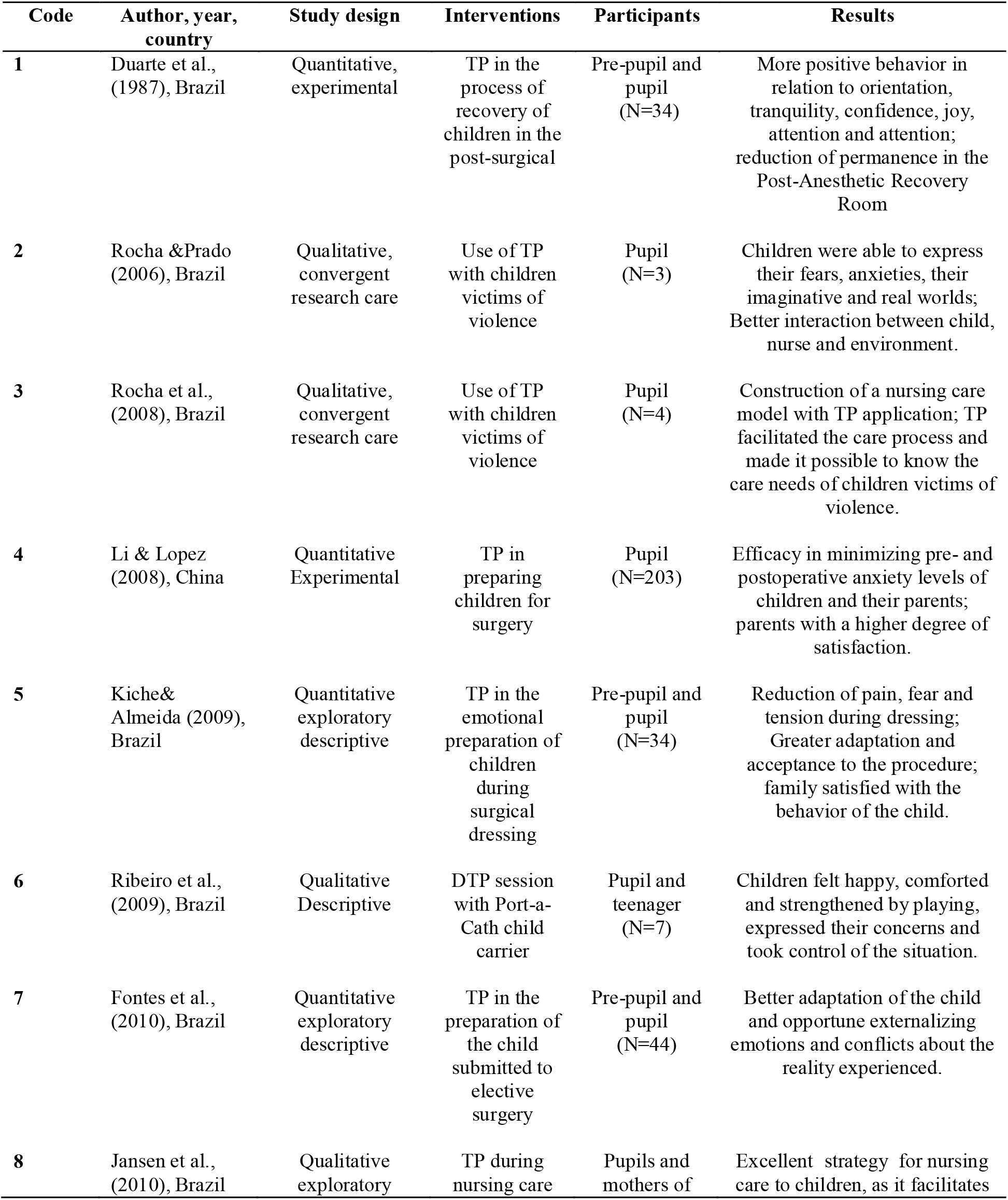
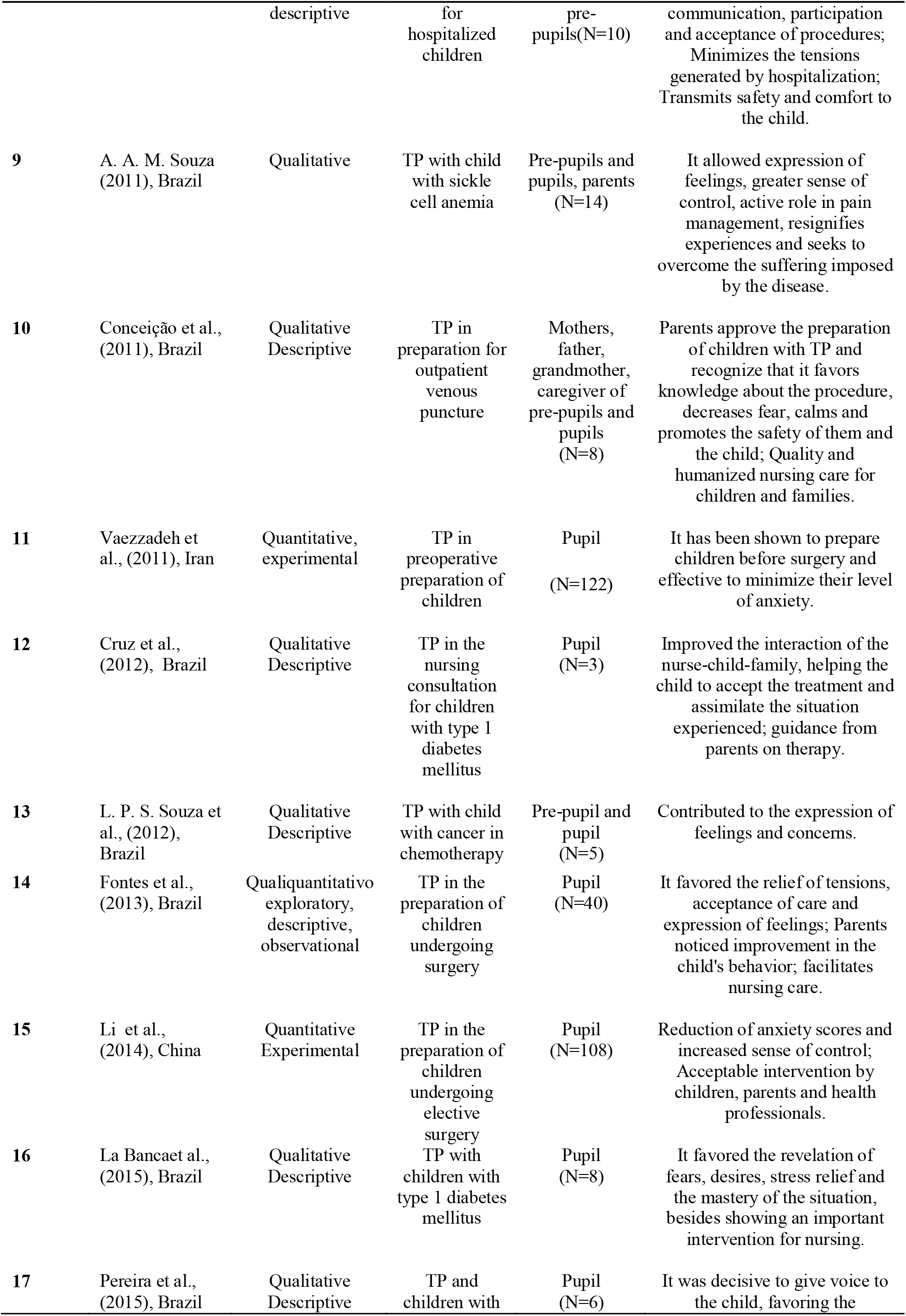
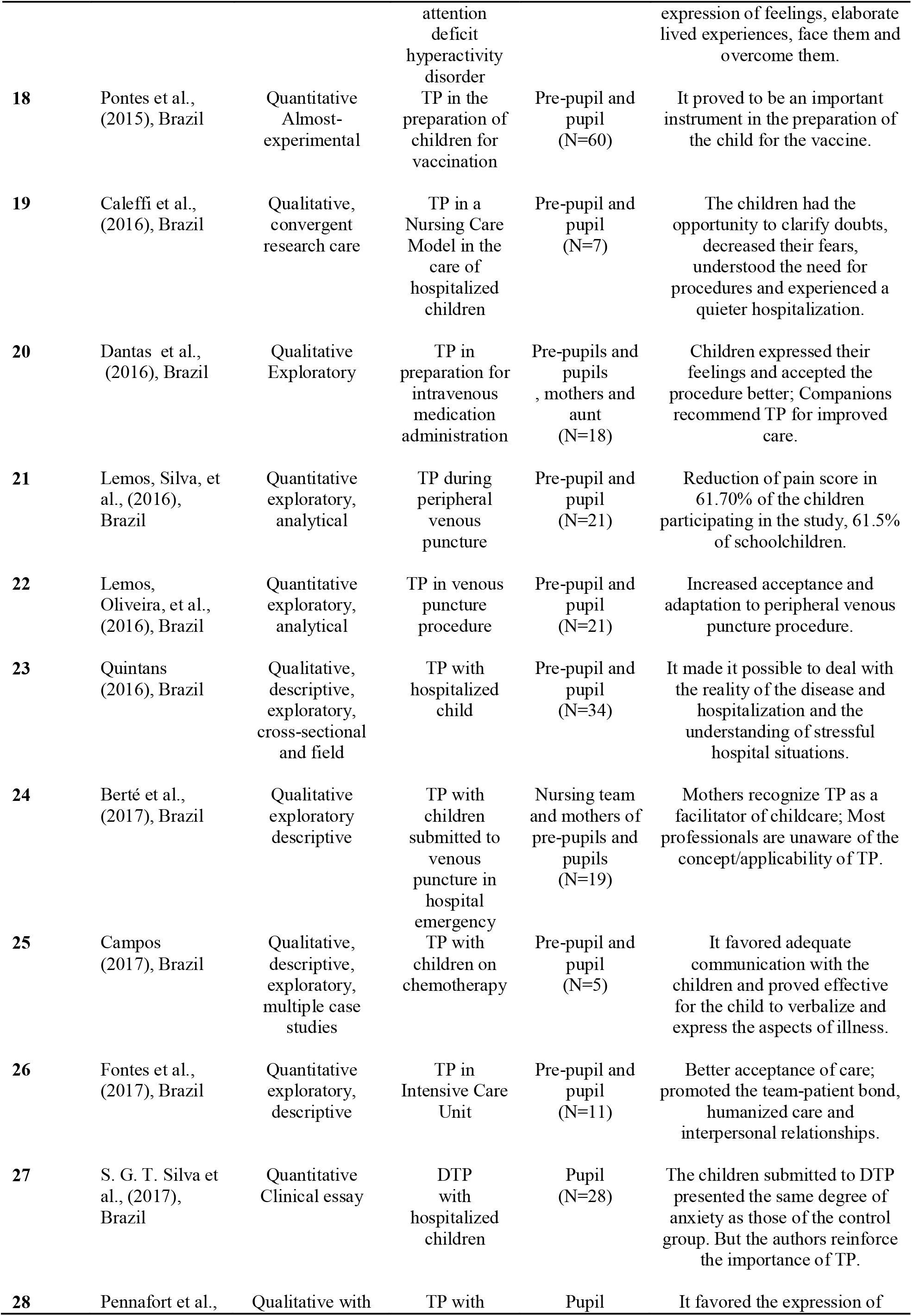
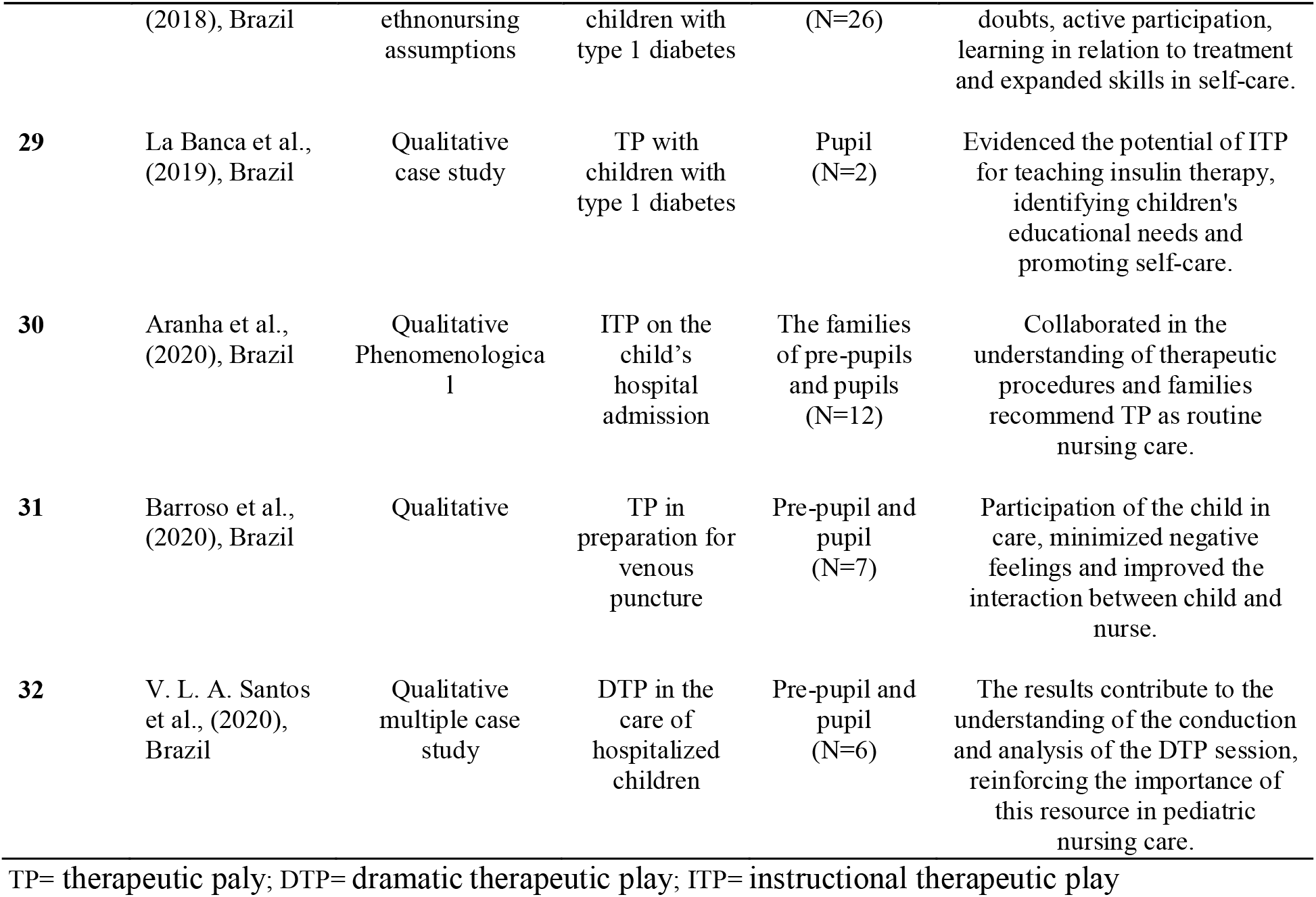
Selected studies in the scoping review on the use of therapeutic play in nursing care for school-age children.

Included studies were published from 1987-2020, with the majority of studies published in 2016 (*n*=5). Among the studies, 29 were original articles, two dissertations and a thesis, available in Portuguese (*n*=18) and In English (*n*=14).

In the methodological design, the studies had qualitative (*n*=20), quantitative (*n*=11) and mixed (*n*=1) approaches. As theoretical references, Erikson’s Theory of Psychosocial Development (*n*=2) (Li & Lopez, 2008; Pereira et al., 2015); Lazarus and Folkman’s theory of cognitive assessment, stress, and coping (*n*=1) (Li & Lopez, 2008); Piaget Cognitive Development Theory (*n*=3) (La Banca et al., 2019; Li & Lopez, 2008; Pereira et al., 2015); Leininger’s Cross-Cultural Theory (*n*=1) (Pennafort et al., 2018); Symbolic Interactionism (*n*=3) (Santos et al., 2020; Souza, 2011; Souza et al., 2012), and Vygotsky’s theory of symbolic child play (*n*=2)(Pereira et al., 2015; Santos et al., 2020).

In general, the studies conducted interventions with TP in order to compare the behavioral reactions of the child before and after TP(*n*=3) and among children who receive and do not receive TP(*n*=2), examine its efficacy(*n*=4), verify its benefits(*n*=7), use it as a therapeutic resource (*n*=5), to understand the experience of the student(*n*=9), analyze the perception of family members/companions and the nursing team about the use of TP (*n*=4) and understand the DTP session (*n*=1).

It is note that 31 studies showed positive outcomes with the use of TP in nursing care for school-age children, corresponding to almost 100% of the review sample.

Fig. 2 presents a diagram with the main findings on the use of TP in nursing care for school-age children.

**Fig. 2.**
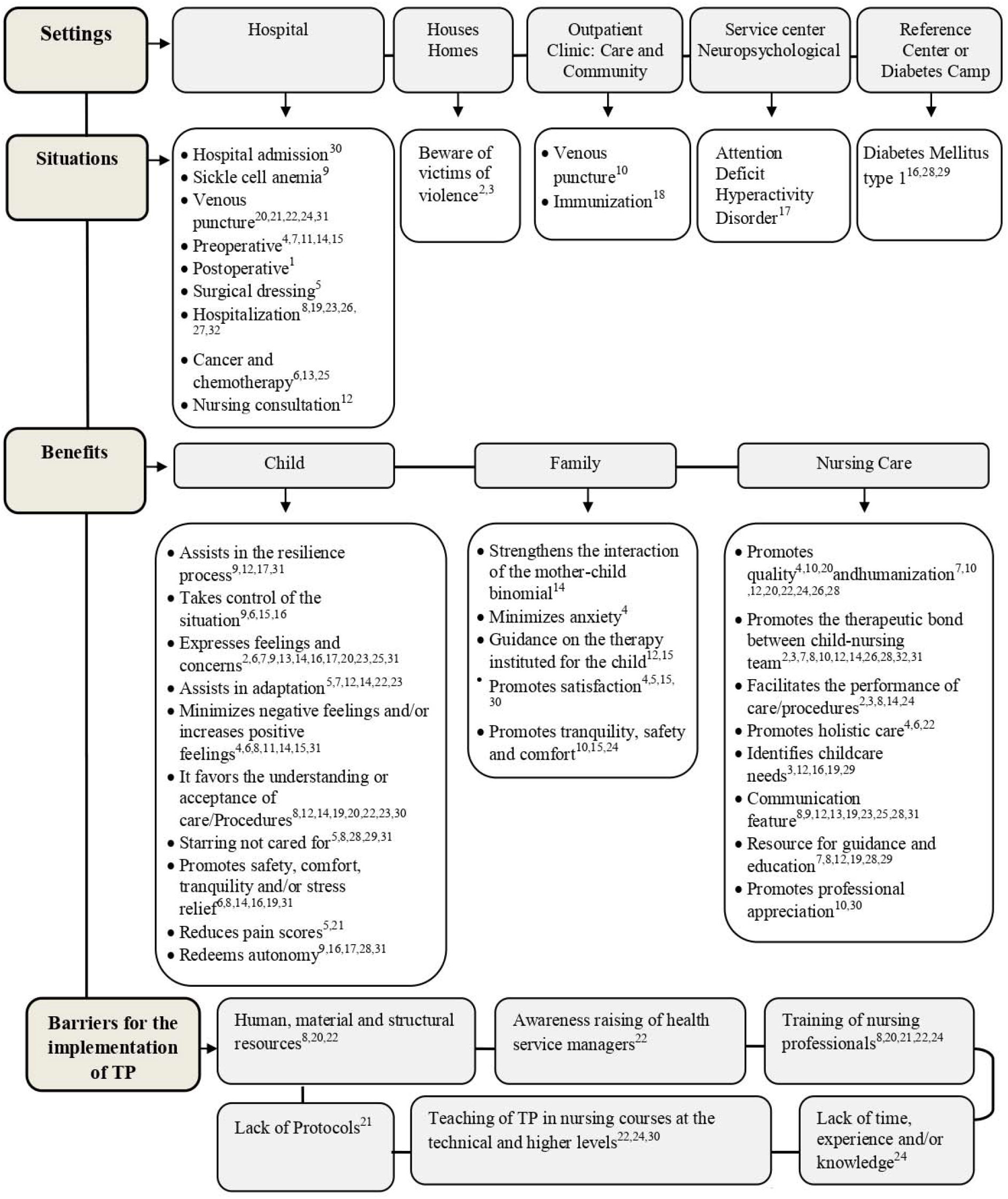
Diagram of the main findings on the use of therapeutic play in nursing care for school-age children.

In the settings of the studies, there was a predominance of the hospital environment in 75% (*n*=24), specifically, in the hospitalization unit (*n*=7), surgical unit (*n*=7), outpatient clinic (*n*=4), clinical (*n*=4), intensive care unit (*n*=2), occupational education and therapy space (*n*=2) and pediatric emergency department (*n*=1). We highlight a study that was conducted in three sectors of one hospital and another study took place in two sectors.

The studies used the ITP and DTP modalities in several situations in nursing care for school-age children. The TP in general was more explored in the hospital environment with care strategy in the hospitalization process (*n*=6), in the preparation for the venous puncture procedure (*n*=5), preoperative (*n*=5) and with children with cancer undergoing chemotherapy treatment (*n*=3).

The ITP involved situations of venous puncture (Berté et al., 2017; Conceição et al., 2011; Dantas et al., 2016; Lemos et al., 2016b), immunization (Pontes et al., 2015), preoperative (Fontes et al., 2010; Fontes et al., 2013; Li et al., 2014; Li &Lopez, 2008; Vaezzadeh et al., 2011), dressing of small and medium-sized surgeries (Fontes et al., 2010; Kiche & Almeida, 2009). Also, during procedures during hospital admission (Aranha et al., 2020), pain management in the face of venous puncture (Lemos et al., 2016a), hospitalization (Caleffi et al., 2016 ; Jansen et al., 2010) and education in glycemic monitoring and insulin therapy for children with diabetes (Cruz et al, 2012; La Banca et al., 2019; Pennafort et al., 2018).

The DTP was used in nursing care in postoperative recovery (Duarte et al., 1987), diabetes mellitus (La Banca et al., 2015), sickle cell anemia (Souza, 2011), Attention Deficit Hyperactivity Disorder (Pereira et al., 2015), accessing a Port-a-Cath and chemotherapy (Campos, 2017; Ribeiro et al., 2009; Souza et al., 2012), venous puncture (Barroso et al., 2020) and hospitalization (Caleffi et al., 2016; Fontes et al., 2017; Quintans, 2016; Santos et al., 2020; S. G. T. Silva et al., 2017), as well as in a nursing consultation with children with type 1 diabetes mellitus (Cruz et al., 2012) and in the construction of a model of nursing care for children victims of violence (sexual abuse, physical abuse or neglect) (Rocha & Prado, 2006; Rocha et al., 2008).

It was found that TP has many benefits, mainly as an ally of the school-age child in the expression of feelings and concerns (*n*=12), favoring understanding or acceptance of care/procedures (*n*=8), increasing positive feelings (satisfaction, joy and motivation) and minimizing negative feelings (pain, anxiety, anguish and stress) (*n*=7) and promoting safety, comfort, tranquility and/or stress relief (*n*=6). For the family, there was a predominance of the feeling of satisfaction (*n*=4), tranquility, safety and comfort (*n*=3) in view of the benefits of TP for their children.

TP can help the nursing professional, being more cited to establish the therapeutic bond with the child (*n*=11), promote communication (*n*=9), humanization (*n*=8) and education/orientation (*n*=6). Among the barriers for the implementation of this strategy in care, we highlight the need for training of nursing professionals (*n*=5), teaching of TP in nursing education at the technical/higher levels (*n*=3) and the need for human, material and structural resources (*n*=3).

## Discussion

The objective of this scoping review was to map the use of therapeutic play in nursing care for school-age children. In this sense, it allowed us to verify that the theme in focus has been discussed over the years in research in the scientific literature. It was identified that the use of TP in nursing care for this age group has been predominantly described in the hospital setting (Aranha et al., 2020; Barroso et al., 2020; Berté et al., 2017; Caleffi et al., 2016; Campos, 2017; Cruz et al., 2012; Dantas et al., 2016; Duarte et al., 1987; Fontes et al., 2010; Fontes et al., 2013; Fontes et al., 2017; Jansen et al., 2010; Kiche & Almeida, 2009; Lemos et al., 2016a; Lemos et al., 2016b; Li et al., 2014; Li & Lopez, 2008; Quintans, 2016; ; Ribeiro et al., 2009; Santos et al., 2020; S. G. T. Silva et al., 2017; Souza, 2011; Souza et al., 2012; Vaezzadeh et al., 2011).

The hospital, in fact, represents a setting in which children can express fear and physical pain due to isolation, lack of engagement with friends and family. Children verbalize and draw about it in some studies around the world, which means it is a universal child reaction. During hospitalization, children face invasive, unknown and painful nursing procedures that can leave them at a high level of anxiety, leading them to express behavior changes (Campos et al., 2020; Cassemiro et al., 2020).

In this context, children’s anxiety can be managed with different clinical actions, such as therapy with games, communication skills and can be measured by different types of tools. Anxiety is an important phenomenon in the field of pediatric nursing care, is defined as a nursing diagnosis and needs to be managed through playful therapy that has positive effects on children’s behavior, pain relief and acceptance of nursing procedures during hospitalization (Campos et al., 2020; Godino-Iáñez et al., 2020; R. D.M. Silva et al., 2017).

It is emphasized that playing is a basic need of every child, even when they are sick, essential for their motor, mental, emotional and social development, being the most genuine way that they use to communicate and express themselves (Paladino et al., 2014). Silva and Cabral (2014) reinforce that the maintenance of play is a need and right of children who demand nursing care of different natures.

The nursing team should be attentive to this need and provide the opportunity of TP for the school-age child, seeking to incorporate into their practices in the health environment whenever appropriate or acceptable (Marques et al., 2015), because TP is an care strategy that can be transferable to all children in different places or cultural contexts (Liet al., 2014).When the toy is provided in a therapeutic way, nursing care becomes a distraction, having a better understanding on the part of the child, allowing greater adaptation and acceptance of therapy. The environment becomes a playful and welcoming atmosphere, which allows the child to express their needs and actively participate in care, softening their doubts, fears and longings (Cruz et al., 2012).

The modalities of ITP and DTP were used in the studies. While there is a gap in the use of ETP, as it was not mentioned in studies identified in this review. The findings show that the importance of ITP and DTP has been reported for different situations, most of them in the hospital context.

Regarding the implementation of the TP sessions, they were performed individually or in groups, being mediated by dolls representative of the family, animals, health professionals, symbolic or real objects of the hospital setting and the domestic setting, with the participation of children handling toys, as well as storytelling illustrating nursing care, followed by dramatization by children. The duration of the sessions ranged from 15 to 60 minutes.

It was noted the lack of protocols to guide the development of TP sessions with the school-age group, highlighting the need for construction and validation of protocols for conducting systematic and effective sessions of the modalities of TP.

A study included in this review (Lemos et al., 2016a) points out that the systematic use of TP in care includes not only the use of regular use, but also the use of validated national and international protocols. In a study (Li et al., 2014) conducted in China, the authors used a protocol for conducting TP when preparing Chinese school-age children preoperatively. A study (Lemos et al., 2016a) conducted in Brazil informs that it has been necessary to adapt existing protocols to use TP in other procedures and other age groups, such as in the study of the authors themselves who used for school-age children a protocol elaborated for preschoolers. They also highlight that, in daily practice, many professionals end up applying the TP sessions in an uncoordinated, irregular and only empirical way.

Given the experience of Martins et al. (2001), the structuring of a protocol for the use of TP with preschoolers contributed to successfully perform the TP technique, was easy to apply and proved effective to achieve the proposed benefits, as well as emphasize that in addition to considering playing as a child’s need in nursing care, it is necessary to consider the characteristics of their development.

In this review, the authors of a study (Caleffi et al., 2016) conducted the systematic application of TP through the Nursing Care Model Caring Playing, which is structured in three stages “Welcoming, Playing and Finishing” and realized that it can be applied to all existing types of TP, with the possibility of being useful beyond the hospital environment.

The importance of TP is undeniable, and the results of this review are perceived as a consensus in the literature about its vast benefits for school-age children, family and nursing, with evidence in studies that highlight its efficacy (Cruz et al., 2012; Lemos et al., 2016a; Li et al., 2014; Li & Lopez, 2008; Vaezzadeh et al., 2011) and show that the family accepts and supports the practice of TP (Aranha et al., 2020; Berté et al., 2017; Conceição et al., 2011; Li et al., 2014; Li & Lopez, 2008).

The studies included in this review reinforce the relevance of integrating the TP in nursing care routine for school-age children and their families, due to the benefits resulting from its use and positive results with the TP for school-age children. Only one study that aimed to evaluate the effect of DTP on the degree of anxiety in hospitalized school-age children submitted to intravenous puncture did not identify a significant reduction in anxiety, but the authors recommend the application of DTP based on other studies that have demonstrated the positive effect in reducing the anxiety of school-age children (S. G. T. Silva et al., 2017).

However, there are still barriers to implement TP in nursing care, such as the need for teaching of TP in nursing training at the technical/higher levels, training of nursing professionals, human resources, materials and structural, lack of protocols, lack of time, experience and/or knowledge of professionals, and sensitization of health service managers (Aranha et al., 2020; Berté et al., 2017; Dantas et al., 2016; Jansen et al.,2010; Lemos et al., 2016a; Lemos et al., 2016b). These barriers need to be overcome and cannot be a hindrance to providing the school-age child with this valuable care strategy that instigates less traumatic care with vast benefits.

To break the barriers of TP implementation, teaching in all nursing training courses at the technical and higher levels is necessary, as it prepares and makes students aware of the application of this strategy in clinical practice (Gesteira et al., 2011). Also, the sensitization of managers of health services and the nursing team about the importance of TP in child health care and care are urgent factors to be considered to effect their daily application by pediatric nursing (Claus et al., 2021).The strategies of continuing education of the nursing team for the use of the ITP, DTP and ETP are relevant, and it is necessary to ensure that these professionals have subsidies of human, structural and material resources for the implementation of this strategy in the routine care of children’s health and to effect child-centered care.

Nurses have been discussing child-centered care (CCC), with increased research on this topic in recent years (Carter & Ford, 2013). In the literature, CCC has been described as an approach in the same terms as person-centered care and family-centered care. In the field of CCC, children are seen as social agents, the nurse’s communication skills are important to recognize the needs, thoughts and actions of children. The CCC proposes to be a means by which nurses can provide a beneficial and therapeutic relationship with the child (Coyne et al., 2018; Ford et al., 2018).

Art-based approaches have been used by nurses to provide care and research with children. Some art-based care options are poetry, music, visual arts, performance, drawing and crafts (Carter & Ford, 2013). Therapeutic play has been reported in different studies and therapeutic play is one of the types that have shown positive effects during childhood hospitalization (Godino-Iáñez et al., 2020; R. D.M. Silva et al., 2017).

A school-age child acquires language and symbolic function, but presents egocentric, playful and magical thinking. For their, the TP represents the opportunity to touch, manipulate, interact in a concrete and symbolic way with dolls and medical devices (such as syringe, gauze and stethoscope, for example) and express their feelings and internal perceptions through play (R. D.M. Silva et al., 2017).

Studies of this review have identified the lack of TP in the routine of nursing care for school-age children (Berté et al., 2017; Dantas et al., 2016; Jansen et al., 2010; Souza et al., 2012). We cannot conclude that this practice in general is routinely scarce, but it was little mentioned in the literature analyzed in this review.

Pediatric nurses should be able to implement care respecting the high standards of care for children and families (Mott et al., 2018). Nursing students demonstrated to receive only theoretical preparation to implement TP sessions, but not practical clinical training during graduation, which left them insecure to conduct the sessions (Barreto et al., 2017; Barroso et al., 2019).

All members of the nursing team play an important role in the promotion and execution of playful activities that help them provide quality and humanized care to the child (Barroso et al., 2020).

Evidence from this review emphasizes that TP is a fast, easy-to-apply and low-cost strategy (Aranha et al., 2020; Li et al., 2014; Li &Lopez, 2008; S. G. T. Silva et al., 2017;) and many (Aranha et al., 2020; Barroso et al., 2020; Campos, 2017; Conceição et al., 2011; Dantas et al., 2016; Jansen et al., 2010; La Banca et al., 2015; Li et al., 2014; Li & Lopez, 2008; Ribeiro et al., 2009; Santos et al., 2020; Souza et al., 2012) promote the awareness of nursing professionals that playing is a fundamental aspect in children’s life and show the importance of TP as an essential strategy for holistic and quality nursing care for children’s health. Therefore, based on the positive results achieved with TP, they encourage integrating it into the planning of pediatric nursing care and be implemented as routine and systematic care strategy whenever appropriate or acceptable for children, as well as applied compatible with the age group.

### Limitations of this study

A possible limitation of this scoping review is related to the number of databases used for bibliographic search, given the possibility of identifying indexed publications in other databases.

## Conclusions

This scoping review identified that the TP is a care strategy with vast benefits for children, families and nursing, but there are still barriers for its implementation in the nursing care routine. There was also that the use of ITP and DTP in nursing care for school-age children has been investigated predominantly in situations related to the hospital setting.

There are gaps regarding the exploration of TP in pediatric nursing practice in out-of-hospital health services. In addition, lack data on the use of ETP and is need for the structuring of protocols to support the conduct of TP sessions with the school-age children.

## Implications for practice

The positive results reported in the studies reinforces the TP as an important care strategy in nursing practice, can be used to enrich the discussions about the use of TP in nursing care for school-age children and encourage awareness about the importance of this resource in care to children.

The gaps identified indicate the need for future research that explores the potential of TP as a nursing care strategy for school-age child in out-of-hospital health services, which generate knowledge about the modality of ETP with the school-age child and the construction of protocols. Favoring expanding scientific knowledge in the area and strengthening the use of TP based on evidence-based practice.

## Data Availability

No data

## References

Aranha, B. F., Souza, M. A., Pedroso, G. E. R., Maia, E. B. S., & Melo, L. L. (2020). Using the instructional therapeutic play during admission of children to hospital: the perception of the family. Revista Gaúcha de Enfermagem, 41, e20180413. https://doi.org/10.1590/1983-1447.2020.20180413

Barreto, L. M. S. C., Maia, E. B. S., Depianti, J. R. B., Melo, L. L., Ohara, C. V. S., & Ribeiro, C. A. (2017). Giving meaning to the teaching of Therapeutic Play: the experience of nursing students. Escola Anna Nery, 21(2), e20170038. https://doi.org/10.5935/1414-8145.20170038

Barroso, M. C. C. S., Machado, M. E. D., Cursino, E. G., Silva, L. R., Depianti, J. R. B., & Silva, L. F. (2019). The therapeutic play in nursing graduation: from theory to practice. Journal of Research Fundamental Care Online, 11(4), 1043–1047. http://doi.org/10.9789/2175-5361.2019.v11i4.1043-1047

Barroso, M. C., Santos, R. S., Santos, A. E., Nunes, M. D., & Lucas, E. A. (2020). Children’s perception of venipuncture through therapeutic toy. Acta Paulista de Enfermagem, 33, 1–8. https://doi.org/10.37689/acta-ape/2020AO0296

Berté, C., Ogradowski, K. R. P., Zagonel, I. P. S., Tonin, L., Favero, L., & Almeida Junior, R. L. (2017). Therapeutic toy in the context of pediatric emergency. Revista Baiana de Enfermagem, 31(3), e20378. http://doi.org/10.18471/rbe.v31i3.20378

Brondani, J. P., & Pedro, E. N. R. (2019). The use of children’s stories in nursing care for the child: an integrative review. Revista Brasileira de Enfermagem, 72(Suppl 3), 333–342. https://doi.org/10.1590/0034-7167-2018-0456

Burns-Nader, S., & Hernandez-Reif, M. (2014). Facilitating Play for Hospitalized Children Through Child Life Services. Children’s Health Care. 45(1), 1–21. http://doi.org/10.1080/02739615.2014.948161

Caleffi, C. C. F., Rocha, P. K., Anders, J. C., Souza, A. I. J., Burciaga, V. B., & Serapião, L. S. (2016). Contribution of structured therapeutic play in a nursing care model for hospitalised children. Revista Gaúcha de Enfermagem, 37(2), e58131. http://doi.org/10.1590/1983-1447.2016.02.58131

Campos, S. M. S. (2017). O brincar de faz de conta de crianças com câncer que se submetem ao processo de quimioterapia [Master’s thesis, Universidade de São Paulo]. Catálogo de teses e dissertações CAPES. https://sucupira.capes.gov.br/sucupira/public/consultas/coleta/trabalhoConclusao/viewTrabalhoConclusao.jsf?popup=true&id_trabalho=5197863

Campos, F. V., Antunes, C. F., Damião, E. B., Rossato, L. M., & Nascimento, L. C. (2020). Anxiety assessment tools in hospitalized children. Acta Paulista de Enfermagem, 33, eAPE20180250. http://doi.org/10.37689/actaape/2020AR02505

Carter, B., & Ford, K. (2013). Researching Children’s Health Experiences: The Place for Participatory, Child-Centered, Arts-Based Approaches. Research in Nursing & Health, 36(1), 95–107. http://doi.org/10.1002/nur.21517

Cassemiro, L. K. D. S., Okido, A. C. C., Furtado, M. C. C., & Lima, R. A. G. (2020). The hospital designed by hospitalized children and adolescents. Revista Brasileira de Enfermagem, 73(Suppl 4), e20190399. http://doi.org/10.1590/0034-7167-2019-0399

Claus, M. I. S., Maia, E. B. S., Oliveira, A. I. B., Ramos, A. L., Dias, P. L. M., & Wernet, M. (2021). The insertion of play and toys in Pediatric Nursing practices: A convergent care research”. Escola Anna Nery, 25(3), e20200383. https://doi.org/10.1590/2177-9465-2020-0383

Conceição, C. M., Ribeiro, C. A., Borba, R. I. H., Ohara, C. V. S., & Andrade, P. R. (2011). Brinquedo terapêutico no prepare da criança para punção venosa ambulatorial: percepção dos pais e acompanhanetes. Escola Anna Nery, 15(2), 346–353. https://doi.org/10.1590/S1414-81452011000200018

Costa, W. M., Sousa, H. O., & Fernandes, M. R. (2019). Brinquedo terapêutico na enfermagem pediátrica brasileira: uma revisão da literatura das evidências atuais. Journal of the Health Sciences Institute, 37(3), 260–263.

Costa, D. T. L., Veríssimo, M. L. O. R., Toriyama, A. T. M., & Sigaud, C. H. S. (2016). O brincar na assistência de enfermagem à criança – revisão integrativa. Revista da Socidade Brasileira de Enfermeiros Pediatras, 16(1), 36–43.

Coyne, I., Holmström, I., & Söderbäck, M. (2018). Centeredness in Healthcare: A Concept Synthesis of Family-centered Care, Person-centered Care and Child-centered Care. Journal of Pediatric Nursing, 42, 45–56. https://doi.org/10.1016/j.pedn.2018.07.001

Cruz, D. S. M., Collet, N., & Marques, D. K. A. (2012). Importance of using therapeutic toys in care of children with diabetes type 1. Journal of Nursing UFPE, 6(4), 849–853. https://doi.org/10.5205/1981-8963-v6i4a7108p849-853-2012

Dantas, F. A., Nóbrega, V. M., Pimenta, E. A. G., & Collet, N. (2016). Use of therapeutic play during intravenous drug administration in children: exploratory study. Online Brazilian Journal of Nursing, 15(3), 1–10. https://doi.org/10.17665/1676-4285.20165581

Duarte, E. R. M., Müller, A. M., Bruno, S. M. A., & Duarte, A. L. S. (1987). A utilização do brinquedo na sala de recuperação: um recurso a mais para assistência de enfermagem à criança. Revista Brasileira de Enfermagem, 40(1), 74–81. https://doi.org/10.1590/S0034-71671987000100013

Fontes, C. M. B., Mondini, C. C. S. D., Moraes, M. C. A. F., Bachega, M. I., & Maximino, N. P. (2010). Utilização do brinquedo terapêutico na assistência à criança hospitalizada. Revista Brasileira de Educação Especial, 16(1), 95–106. https://doi.org/10.1590/S1413-65382010000100008

Fontes, C. M. B., Oliveira, A. S. S., & Toso, L. A. (2017). Brinquedo terapêutico em unidade de terapia intensiva pediátrica. Revista de Enfermagem UFPE, 11(Supl.7), 2907–2915. https://doi.org/10.5205/reuol.11007-98133-3-SM.1107sup201712

Fontes, C. M. B., Sá, F. M., Mondini, C. C. S. D., & Moraes, M. C. A. F. (2013). Therapeutic toy and child preparation for correction of cleft lip and palate surgery. Journal of Nursing UFPE, 7(7), 4681–4688. https://doi.org/10.5205/reuol.4656-38001-2-SM.0707201313

Ford, K., Campbell, S., Carter, B., & Earwaker, L. (2018). The concept of child-centered care in healthcare: a scoping review protocol. JBI Database of Systematic Reviews and Implementation Reports, 16(4), 845–851. https://doi.org/10.11124/JBISRIR-2017-003464

Francischinelli, A. G. B., Almeida, F. A., & Fernandes, D. M. S. O. (2012). Routine use of therapeutic play in the care of hospitalized children: nurses’ perceptions. Acta Paulista de Enfermagem, 25(1), 18–23. https://doi.org/10.1590/S0103-21002012000100004

Gesteira, E. R., Gonçalves, D. S., Marques, F., & Simões, F. D. (2011). A experiência de alunos na utilização do brinquedo terapêutico no estágio de enfermagem pediátrica. Revista de Enfermagem da UFPE, 5(7), 1807–1811. https://doi.org/10.5205/reuol.1262-12560-1-LE.0507201132

Godino-Iáñez, M. J., Martos-Cabrera, M. B., & Suleiman-Martos, N., Urquiza, J. L. G., Vargas-Román, K., Membrive, M. J., & Albendin, L. (2020). Play Therapy as an Intervention in Hospitalized Children: A Systematic Review. Healthcare, 8(3), 239. https://doi.org/10.3390/healthcare8030239

Hockenberry, M. J, & Wilson, D. (2011). Wong: fundamentos de enfermagem pediátrica. Elsevier..

Jansen, M. F., Santos, R. M., & Favero, L. (2010). Benefícios da utilização do brinquedo durante o cuidado de enfermagem prestado a criança hospitalizada. Revista Gaucha de Enfermagem, 31(2), 247–253. https://doi.org/10.1590/S1983-14472010000200007

Kiche, M. T., & Almeida, F. A. (2009). Therapeutic toy: strategy for pain management and tension relief during dressing change in children. Acta Paulista de Enfermagem, 22(2), 125–130. https://doi.org/10.1590/S0103-21002009000200002

La Banca, R. O., Monteiro, O. O., Ribeiro, C. A., & Borba, R. I. H. (2015). School experience of children with diabetes mellitus expressed by dramatic therapeutic play. Journal of Nursing UFPE, 9(Supl. 7), 9009–9017. https://doi.org/10.5205/reuol.8074-70954-1-SM0907supl201510

Lemos, I. C. S., Oliveira, J. D., Gomes, E. B., Silva, K. V. L., Silva, P. K. S., & Fernandes, G. P. (2016b). Brinquedo terapêutico no procedimento de ounção venosa: estratégia para reduzir alterações comportamentais. Revista Cuidarte, 7(1), 1163–1170. https://doi.org/10.15649/cuidarte.v7i1.303

Lemos, I. C. S., Silva, L. G., Delmondes, G. A., Brasil, A. X., Santos, P. L. F., Gomes, E. B., Silva, K. V. L. G., Oliveira, D. R., Oliveira, J. D., Fernandes, G. P., & Kerntopf, M. R. (2016a). Therapeutic Play Use in Children under the Venipucture: A Strategy for Pain Reduction. American Journal of Nursing Research, 4(1), 1–5. https://doi.org/10.12691/ajnr-4-1-1

Li, H. C., & Lopez, V. (2008). Effectiveness and appropriateness of therapeutic play intervention in preparing children for surgery: a randomized controlled trial study. Journal for Specialists in Pediatric Nursing, 13(2), 63–73. https://doi.org/10.1111/j.1744-6155.2008.00138.x

Li, W. H. C., Chan, S. S. C., Wong, E. M. L., Kwok, M. C., & Lee, I. T. L. (2014). Effect of therapeutic play on pre- and post-operative anxiety and emotional responses in Hong Kong Chinese children: a randomised controlled trial. Hong Kong Medical Journal, 20(Suppl 7), 36–39.

Maia, E. B. S., Melo, L. L., La Banca, R. O. (2020). A criança, o adolescente e a hospitalização. In: N. Collet, B. R. G. O. Toso, & C. S. Vieira (Eds.), Manual de Enfermagem em pediatria (pp. 49–53). AB editora.

Marques, D. K. A., Silva, K. L. B., Cruz, D. S. M., & Souza, I. V. B. (2015). Benefícios da aplicação do brinquedo terapêutico: visão dos enfermeiros de um hospital infantil. Arquivos de Ciências da Saúde, 22(3), 64–68. https://doi.org/10.17696/2318-3691.22.3.2015.240

Martins, M. R., Ribeiro, C. A., Borba, R. I. H., & Silva, C. V. (2001). Protocolo de preparo da criança pré-escolar para punção venosa, com utilização do brinquedo terapêutico. Revista Latino-Americana de Enfermagem, 9(2), 76–85. https://doi.org/10.1590/S0104-11692001000200011

Melo, L. L., Ribeiro, C. A. (2020). (Cre)scendo na ausência da mãe: vivências de crianças durante o cárcere materno. Revista Brasileira de Enfermagem, 73(Suppl 4), e20200413. https://doi.org/10.1590/0034-7167-2020-0413

Moher. D., Liberati, A., Tetzlaff, J., & Altman, D. G. (2009). Preferred reporting items for systematic reviews and meta-analyses: the PRISMA statement. PloS Medicine, 6(7), e1000097. https://doi.org/10.1371/journal.pmed.1000097

Mott, S., Fogg, N., Foote, N., Hillier, M., Lewis, D. A., Mcdowell, B. M., Saunders, K., Taylor, J. T., Wiggins, S., Ivey, J. B., Benedetto, C. O., Beam, P. H., Mcknight, K. B., Taha, A. A., & Vann-Patterson, A. (2018). Society of Pediatric Nurses’ Core Competencies for the Pediatric Nurse. Journal of Pediatric Nursing, 38, 142–144. https://doi.org/10.1016/j.pedn.2017.11.006

Nóbrega, R. D., Collet, N., Gomes, I. P., Holanda, E. R., & Araújo, Y. B. (2010). Criança em idade escolar hospitalizada: significado da condição crônica. Texto & Contexto Enfermagem, 19(3), 425–433. https://doi.org/10.1590/S0104-07072010000300003

Oliveira, C. S., Maia, E. B. S., Borba, R. I. H., & Ribeiro, C. A. (2015). Brinquedo Terapêutico na assistência à criança: percepção de enfermeiros das unidades pediátricas de um hospital universitário. Revista da Sociedade Brasileira de Enfermeiros Pediatras, 15(1), 21–30.

Paladino, C. M., Carvalho, R., & Almeida, F. A. (2014). Therapeutic play in preparing for surgery: behavior of preschool children during the perioperative period. Revista da Escola de Enfermagem da USP, 48(3), 423–429. https://doi.org/10.1590/S0080-623420140000300006

Pennafort, V. P. S., Queiroz, M. V. O., Gomes, I. L. V., & Rocha, M. F. F. (2018). Instructional therapeutic toy in the culture care of the child with diabetes type 1. Revista Brasileira de Enfermagem, 71(Suppl 3), 1334–1342. https://doi.org/10.1590/0034-7167-2017-0260

Pereira, A. K., Maia, E. B. S., Borba, R. I. H., & Ribeiro, C. A. (2015). O brincar da criança com transtorno de déficit de atenção e hiperatividade. Ciência, Cuidado e Saúde, 14(2), 1175–1183.

Peters, M. D. J., Godfrey, C., McInerney, P., Munn, Z., Tricco, A. C., & Khalil, H. (2020). Scoping Reviews (2020 version). In: E. Aromataris & Z. Munn (Eds), Joanna Briggs Institute Reviewer’s Manual. The Joanna Briggs Institute. https://reviewersmanual.joannabriggs.org/.

Pontes, J. E. D., Tabet, E., Folkmann, M. A. S., Cunha, M. L. R., & Almeida, F. A. (2015). Therapeutic play: preparing the child for the vaccine. Einstein, 13(2), 238–242. https://doi.org/10.1590/S1679-45082015AO2967

Quintans, D. E. B. (2016). O jogo ‘faz de conta” na sessão de brinquedo terapêutico de crianças hospitalizadas [Master’s thesis, Universidade Guarulhos]. Catálogo de teses e dissertações CAPES. https://sucupira.capes.gov.br/sucupira/public/consultas/coleta/trabalhoConclusao/viewTrabalhoConclusao.jsf?popup=true&id_trabalho=4055876

Ribeiro, C. A., Coutinho, R. M., Araújo, T. F., & Souza, V. S. (2009). A world of procedures and worries: Experience of children with a Port-a-Cath. Acta Paulista de Enfermagem, 22(spe), 935–941. https://doi.org/10.1590/S0103-21002009000700017

Rocha, P. K., & Prado, M. L. (2006). Violência infantil e brinquedo terapêutico. Revista Gaúcha de Enfermagem, 27(3), 463–471.

Rocha, P. K., Prado, M. L., & Carraro, T. E. (2008). Nursing care model for children victims of violence. Australian Journal of Advanced Nursing, 25(3), 80–85.

Santos, V. L. A., Almeida, F. A., Ceribelli, C., & Ribeiro, C. A. (2020). Understanding the dramatic therapeutic play session: a contribution to pediatric nursing. Revista Brasileira de Enfermagem, 73(4), e20180812. https://doi.org/10.1590/0034-7167-2018-0812

Santos, F. S., Pereira, Y. J. A. S., Sá, M. C. N., Costa, A. C. P. J., Fontoura, I. G., Serra, M. A. A. O., Santos Neto, M., Ferreira, A. G. N., Noleto, A. A. S., & Santos, F. D. R. P. (2017). Nursing intervention with hospitalized children mediated by therapeutic toys. International Journal of Development Research, 7(10), 15821–15826.

Silva, R. D. M., Austregésilo, S. C., Ithamar, L., & Lima, L. S. (2017). Therapeutic play to prepare children for invasive procedures: a systematic review. Jornal de Pediatria, 93(1), 6–16. https://doi.org/10.1016/j.jped.2016.06.005

Silva, L. F., & Cabral, I. E. (2014). Cancer repercussions on play in children: implications for nursing care. Texto Contexto Enfermagem, 23(4), 935–943. https://doi.org/10.1590/0104-07072014002380013

Silva, S. G. T., Santos, M. A., Floriano, C. M. F., Damião, E. B. C., Campos, F. V., & Rossato, L. M. (2017). Influence of Therapeutic Play on the anxiety of hospitalized school-age children: Clinical trial. Revista Brasileira de Enfermagem, 70(6), 1244–1249. https://doi.org/10.1590/0034-7167-2016-0353

Souza, A. A. M. (2011). Uma vida dominada pela dor: a criança vivenciando a anemia falciforme. [Master’s thesis, Universidade Federal de São Paulo]. Repositório Institucional da Universidade Federal de São Paulo. https://repositorio.unifesp.br/xmlui/handle/11600/10153?locale-attribute=pt_BR

Souza, L. P. S., Silva, R. K. P., Amaral, R. G., Souza, A. A. M., Mota, E. C., & Silva, C. S. O. (2012). Câncer infantil: sentimentos manifestados por crianças em quimioterapia durante sessões de brinquedo terapêutico. Revista Rene, 13(3), 686–692.

The United Nations Children’s Fund. (1990). Convention on the Rights of the Child text. [cited 2020 Apr 23]. Available from: https://www.unicef.org/child-rights-convention/convention-text

Tricco, A. C., Lillie, E., Zarin, W., O’Brien, K. K., Colquhoun, H., Levac, D., …Straus, S. E.. (2018). PRISMA Extension for Scoping Reviews (PRISMAScR): Checklist and Explanation. Annals of Internal Medicine,169(7), 467–73. https://doi.org/10.7326/M18-0850

Vaezzadeh, N., Douki, Z. E., Hadipour, A., Osia, S., Shahmohammadi, S., Sadeghi, R. (2011). The effect of performing preoperative preparation program on school age children’s anxiety. Iran Journal Pediatr[Internet]. [cited 2020 Jul 5];21(4):461–66.

World Health Organization Regional Office for Europe (2017). Children’s rights in Hospital: Rapid-assessment checklists. http://www.euro.who.int/pubrequest.

World Health Organization (2018). Standards for improving the quality of care for children and young adolescents in health facilities. https://www.who.int/maternal_child_adolescent/documents/quality-standards-child-adolescent/en/.

